# Delay of molecular SARS-CoV-2 testing and turnaround time in Mexico and Colombia

**DOI:** 10.1101/2021.06.16.21259046

**Authors:** Isaac Núñez, Pablo F. Belaunzarán-Zamudio, Yanink Caro-Vega

**Author notes:** **Corresponding author**: Isaac Núñez MD. Instituto Nacional de Ciencias Médicas y Nutrición Salvador Zubirán, Vasco de Quiroga #15, Tlalpan Mexico City, Mexico, postal code 14080.

## Abstract

**Objective:** To quantify the delay in SARS-CoV-2 real time polymerase chain reaction (RT-PCR) testing and test result reporting in Mexico and Colombia, and to assess the relation between margination status and these delays.

**Methods:** We quantified time in days from symptom onset until testing (latency one) and delay in test results report (latency two) using freely available country-wide open data from Mexico and Colombia. Directed acyclic graphs were built to determine which associations were appropriate to assess. Stratification by margination status, state and hospitalization status was used to determine corresponding associations.

**Results:** In almost all the study period latency two was longer than latency one. Median latency one was 3 (IQR 0-6) days and latency two 7 (IQR 4-11) days in Colombia, while in Mexico they were 3 (IQR 1-5) days and 4 (IQR 3-6) days. In Colombia, worse margination status prolonged latency two. In Mexico, a lower number and percentage of point-of-care (POC) tests in areas with worse margination.

**Conclusion:** POC tests must be used as a widespread means to reduce latency two, and until then should be prioritized in areas with longer latency two. An unequal distribution of this resource should be avoided.

## Introduction

Since the COVID-19 pandemic started, massive testing has been a staple of epidemic control worldwide to detect cases, initiate contact tracing and quarantine individuals who were exposed (*1*). Real-time reverse-transcriptase polymerase chain reaction (RT-PCR) was the first available test and remains widely utilized (*2, 3*). Being the only test available at the beginning of the pandemic, it was used both for epidemiologic surveillance and individual diagnosis (*4*). It has an almost perfect specificity but suboptimal sensitivity, making false negative results a relevant problem in high prevalence areas, as it becomes easy to undercount cases and misinterpret a negative result (*5*).

An important issue is the timing of the tests, and there are two main reasons for this. If an RT-PCR test is performed late during the natural history of the infection, it could be futile for means of epidemiologic surveillance (*6, 7*). A person that is tested in their 5^th^ day of symptoms but obtained their result 5 days later has remained without a diagnosis for 10 symptomatic days, which could impact their ability (if their job required a positive test to justify absence) or willingness to isolate (*7*). If they tested positive in their 20^th^ day from symptom onset they have ceased to be contagious, and the test would only help to determine if they were infected (and even then, the sensitivity would be suboptimal for that purpose and an antibody assay could be more useful) (*8*). Conversely, a positive test in a person who is on their 5^th^ day of symptoms would allow for relatively early isolation and contact tracing. Thus, delay in test results report has a negative impact both in epidemiological surveillance and individual diagnosis. Using open data, we describe the frequency and length of delays in SARS-CoV-2 RT-PCR tests after symptom initiation and in laboratory tests results report and determine its association with margination status.

## Methods

We searched in the Our World in Data COVID-19 pandemic coverage, an initiative by the University of Oxford, for COVID-19 open access data that met eligibility criteria for our study (*3*). We selected open access country datasets, available in a machine-readable format (e.g. csv, xsl, json, etc) that were regularly updated (at least weekly), and containing at least the following variables at the individual level: date of symptoms onset, date of COVID-19 testing, date of tests result report, and type of test used for diagnosis (RT-PCR, antigen, clinical diagnosis, other). If information on the type of test used for diagnosis was not available (either included as a variable or in the dataset description), we included the country if we found a description in which test use was described (for example, if a statement of the country’s health authority said that antigen tests were introduced at a given date). We excluded datasets of countries with no available information on type of test used for diagnosis and date. For date of testing and date of diagnosis, countries were also eligible if these variables could be calculated with information provided in the dataset. No language restriction was used, and Google translate was used when no English or Spanish version of a given website was available. As RT-PCR is considered the gold standard, we excluded countries that did not use this type of tests or had no information on the kind of test used. Study period could differ between databases. We considered the date in which an individual was first included to be the beginning of the study period. Individual patients were included until January 31^st^ 2021, and final follow up was February 10^th^ 2021 to allow for delayed reporting.

### Definitions

We were interested in exploring delays at two steps of the diagnostic process: delays in seeking care by individuals and delays in the health system in reporting diagnostic test reports. Thus, we generated variables to analyze these as outcomes. Diagnosis date was established searching daily databases using a loop for the date in which a result was first reported for any given individual. We defined *latency one* as the time in days between date of symptom onset and the date of testing. We assume this period is mainly determined by an individual’s characteristics. *Latency two* was defined as the period of time in days between the date of sample collection and the date of test results reporting and we assume this period was determined by factors related to the health systems. We considered the total time between date of symptom onset and date of test result, defined as *total latency*, as the sum of both latencies for each individual.

We classified individuals as *early testers* if they were tested for COVID-19 within the first 3 days of symptom onset, *late testers* if tested between the fourth and eighth day of symptom onset, and *very late testers* if tested after the eighth day of symptom onset. We also classified the second latency period as *efficient* if the RT-PCR result was available within two days of sampling, and *inefficient* if it took longer. Finally, *total latency* was classified as *optimal* if it was shorter than 5 days, *regular* if it was between 6 and 10 days long, or *inadequate* if it was more than 10 days. A recent systematic review and meta-analysis reports that virus transmissibility begins two days before symptoms onset up to 9 days after tit (*11*). Thus, we used this interval of time (extending until 10 days after symptom started for practicality) to define patients as “infectious”.

### Statistical analysis

We used rolling means to describe the length of latencies *one, two*, and *total latency* during the study period, and proportions to describe the frequency of early, late and very late testers (*latency one*); *efficient* and *inefficient* test results reporting (*latency two)*, and *optimal, regular, inadequate total latency*. We calculated the amount of time that would be saved with point-of-care tests, which would eliminate latency two, dividing the total amount of latency two and the number of tested patients. We stratified the analysis by margination status, which was calculated differently for Colombia and Mexico. For Colombia we included the “multidimensional poverty index” (MPI), which is calculated yearly using a national representative household census (*12, 13*). It evaluates five key aspects: education, childhood and youth, health, work, and living place (*13*). A score of zero (no deprivation) and one (deprivation) is then calculated for each household and the proportion of deprived households sampled by country and state (the smallest specified geographical area) is then reported. The most recent version corresponds to the 2019 census (*14*). For Mexico, we calculated the “margination index” (MI), a metric developed by the Mexican Consejo Nacional de Población (National Population Council, CONAPO) also calculated for each household sampled in a national census. The smallest specified geographic area for which this index is calculated is a “locality”. It is constructed by the percentage of people that have each of nine socio-economic characteristics for that geographical area: older than 15 years and illiterate, older than 15 years and incomplete elementary school, no sewer system, no electricity, no tubing water, household overcrowding, dirt floor, less than 5,000 habitants in the locality, and population with an income of maximum the equivalent to two minimum wages. This last variable is not available in the most recent census, so we will substitute it for “percentage of people above age 12 that are not economically active or currently attending school”. Each variable weights 1/9, and with a score that ranges from 0 to 100 (higher score indicating higher margination). The 2020 census did not include the MI, so it was manually calculated using the information on individual variables with the open database created by the National Institute of Statistics and Geography (INEGI, due to its initials in Spanish). In the COVID-19 open database each individual is identified up to municipality (multiple localities join to form a municipality), so a mean score was calculated for each municipality using the score of the localities that conform each municipality, using the total population of each locality as a weight. We stratified patients by MPI or MI quintiles.

To determine whether confounding could impair the validity of stratification by multidimensional poverty index /margination index, we constructed two directed acyclic graphs (DAGs), one for each latency period as the outcome and multidimensional poverty index /margination index as the main exposure (**Appendix Figure 1**). In these causal models, multidimensional poverty index /margination index were considered as the same variable. The proportion of point-of-care tests across margination status was calculated for Mexico to determine accessibility to said tests beginning in November 15^th^, 2020, the date in which these tests started to be performed. Colombia did not have the data to make this calculation.

All analyses were done with R version 4.0.0. DAGs were built with the *daggitty* open software (*15*). Data used in the analysis is freely available in respective government websites (9, *10*), and code used in the analysis will be made available in the final version. The study was approved by the ethics board of the Instituto Nacional de Ciencias Médicas y Nutrición Salvador Zubirán.

## Results

### Characteristics of datasets

Only datasets from Mexico and Colombia met inclusion criteria (*9, 10*). Open data included different information for each country. Only confirmed cases were available for Colombia, while all tested people were available for Mexico. People were registered in the Colombian database from March 2^nd^ 2020, and the study period had to be limited until July 27^th^ because antigen tests began to be reported starting July 28^th^ with no distinction between positive cases by RT-PCR or antigen tests. Mexico does not provide a testing nor a diagnosis date. Testing date was assumed to be that in which an individual first appeared in a database. As these individuals are included with a “pending result” in the result variable, we consider this to be reasonable.

Colombia had a total of 327,366 reported positive individuals from April 1^st^ until August 9^th^ (date in which the last positive test taken during the period in which only RT-PCR were performed was reported) and Mexico had 3,363,456 RT-PCR tested individuals from April 12^th^ 2020 until January 31^st^ 2021 until, of which 3,214,281 had an available result by February 10^th^ and 151,671 had more than 10 days since their test was taken and no available result.

### Time between symptoms onset and testing (latency one)

According to our DAG (**Appendix Figure 1**), the total effect of multidimensional poverty index /margination index on latency one or latency two can be directly estimated by stratification on this variable as the backdoor criterion is satisfied (*15*).

In Colombia, median latency one was 3 days long (IQR 0-6), latency two 7 days (IQR 4-11), and total latency 12 days (IQR 7-15). In Mexico the corresponding values were 3 days (IQR 1-5), 4 days (IQR 3-6), and 8 days (5-12). Latency periods by country and margination quintiles are shown in Table 1. Over time, *latency one* remained stable during the study period in Mexico. In Colombia, it had an initial peak at the beginning of the study period but decreased soon and remained stable time in Colombia (**Figure 1**). Length of latency one was similar across margination status in both countries while latency two was markedly higher in places with worse margination in Colombia, but not in Mexico (**Table 1, Appendix Figure 2**).

**Table 1.** Latency periods of RT-PCR for SARS-CoV-2 testing (*latency one)* and test results report (*latency two*)

| Variable | Colombia | Mexico |
| --- | --- | --- |
| **Latency period in days<sup>1</sup>, median (IQR)** | - | - |
| Latency one | 3 (0-6) | 3 (1-5) |
| Latency two | 7 (4-11) | 4 (3-6) |
| Total latency | 12 (7-15) | 8 (5-12) |
| **Type of tester by day of test<sup>2</sup>, n (%):** | - | - |
| Early testers | 242,965 (74.2) | 1,861,026 (55.3) |
| Late testers | 67,383 (18.8) | 1,238,593 (36.8) |
| Very late testers | 23,018 (7) | 263,788 (7.9) |
| **Efficient test results report<sup>3</sup>, n (%)** | 41,497 (12.6) | 691,030 (20.5) |
| **Classification of total latency, n (%):** | - | - |
| Optimal | 46,799 (14.3) | 815,504 (24.2) |
| Regular | 91,796 (28) | 1,407,143 (41.8) |
| Inadequate | 188,771 (57.7) | 975,492 (29) |
| **Latency by margination status<sup>5</sup>** | - | - |
| First quintile (lowest margination) |  |  |
| -Median latency one (IQR) | 3 (0-7) | 3 (1-5) |
| -Median latency two (IQR) | 7 (4-10) | 5 (3-8) |
| -Median total latency (IQR) | 11 (7-16) | 8 (6-13) |
| Second quintile |  |  |
| -Median latency one (IQR) | 4 (1-6) | 3 (1-5) |
| -Median latency two (IQR) | 6 (4-11) | 4 (3-6) |
| -Median total latency (IQR) | 11 (7-15) | 8 (5-11) |
| Third quintile |  |  |
| -Median latency one (IQR) | 3 (0-5) | 3 (1-5) |
| -Median latency two (IQR) | 7 (4-11) | 4 (3-7) |
| -Median total latency (IQR) | 11 (7-15) | 8 (5-12) |
| Fourth quintile |  |  |
| -Median latency one (IQR) | 4 (1-5) | 3 (2-5) |
| -Median latency two (IQR) | 8 (4-11) | 4 (3-6) |
| -Median total latency (IQR) | 12 (8-15) | 8 (6-11) |
| Fifth quintile (highest margination) |  |  |
| -Median latency one (IQR) | 3 (0-5) | 3 (2-5) |
| -Median latency two (IQR) | 10 (6-11) | 4 (3-6) |
| -Median total latency (IQR) | 14 (9-15) | 8 (6-11) |

<sup>1</sup>Latency one: period from symptom onset until testing; Latency two: period from testing until result availability; Total latency: time from symptom onset until result availability.
<sup>2</sup>“Early testers” were tested within the first three days of symptoms, “late testers” from the fourth day until the eighth day, and “very late testers” afterwards.
<sup>3</sup>Defined as those that were reported within two days from testing date.
<sup>4</sup>Total latency was classified as “optimal” if it was shorter than 5 days, “regular” if it was between 6 and 10 days long, or “inadequate” if it was more than 10 days.
<sup>5</sup>Multidimensional poverty index for Colombia, Margination index for Mexico.

**Figure 1.**
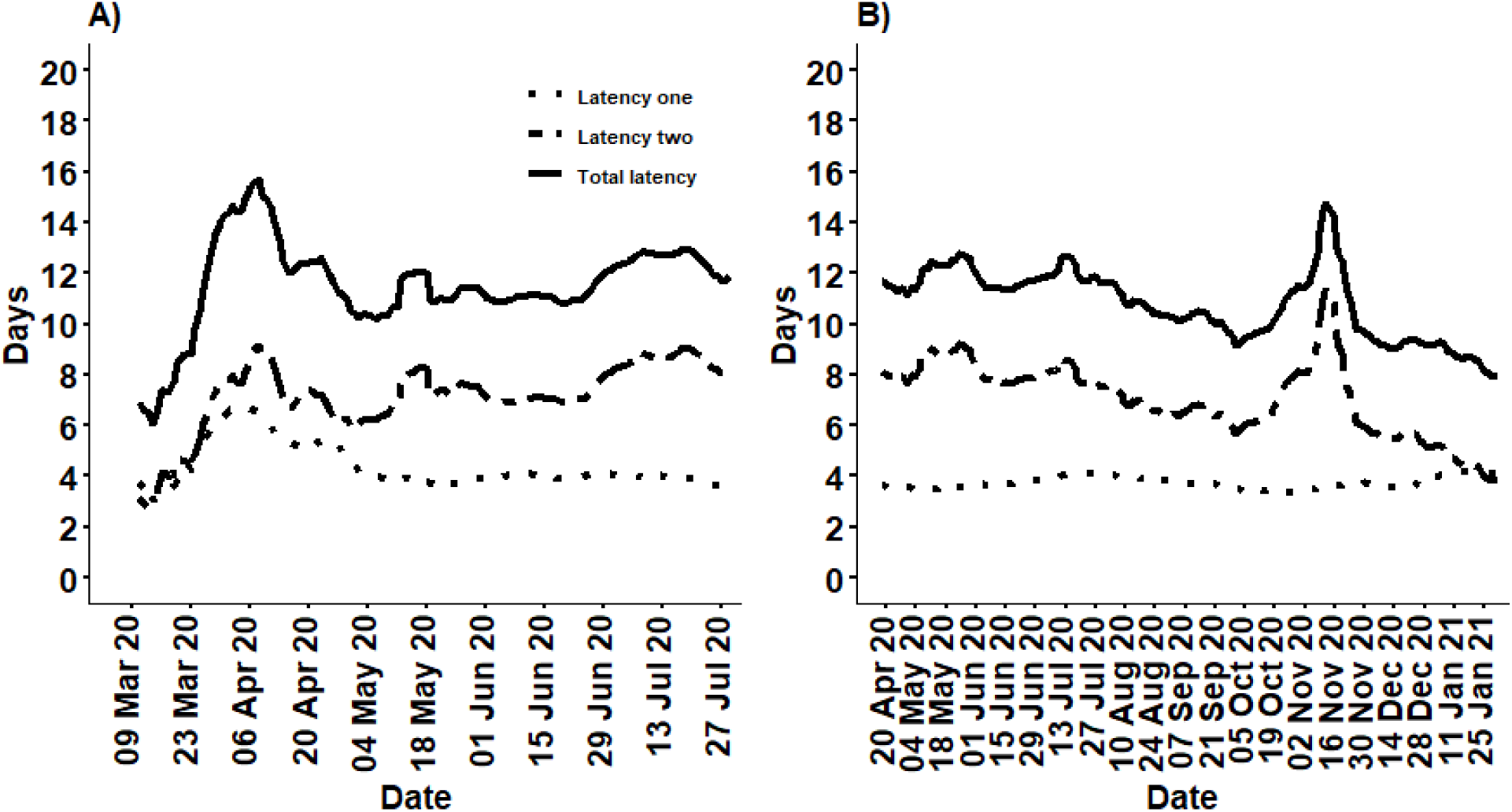
A) Colombia, B) Mexico; Latency one: time from first symptom until testing; Latency two: time from testing until reporting of result; Total latency: total time from first symptom until reporting of test result.

### Time between testing and test result report (latency two)

*Latency two* was much longer than *latency one* in both countries, causing the lengthiest delay in tests result reports. While there was a tendency towards the reduction of *latency two* in Mexico over time (interrupted by a huge peak in October-November 2020), the length of *latency two* increase over time in Colombia (**Figure 1**). We observed a clear tendency towards a lengthier *latency two* in the lowest quintiles of margination status indicators in Colombia but not in Mexico. The number and proportion of performed tests that were point-of-care in Mexico was lower in the worst margination quintiles across the study period, even if by the end the percentage was similar across quintiles (**Table 2, Figure 2**).

**Table 2.** Point-of-care tests according to margination status quintiles in Mexico.

| Margination status | Total tests | POC tests (%) |
| --- | --- | --- |
| Quintile 1 | 474,311 | 302,431 (63.8) |
| Quintile 2 | 531,135 | 355,065 (66.9) |
| Quintile 3 | 508,993 | 345,083 (67.8) |
| Quintile 4 | 368,728 | 144,461 (39.2) |
| Quintile 5 | 335,448 | 81,661 (24.3) |

POC: point-of-care. Fifth quintile represents the worst margination status. Study period for this
calculation is from November 15<sup>th</sup> – January 31<sup>st</sup>.

**Figure 2.**
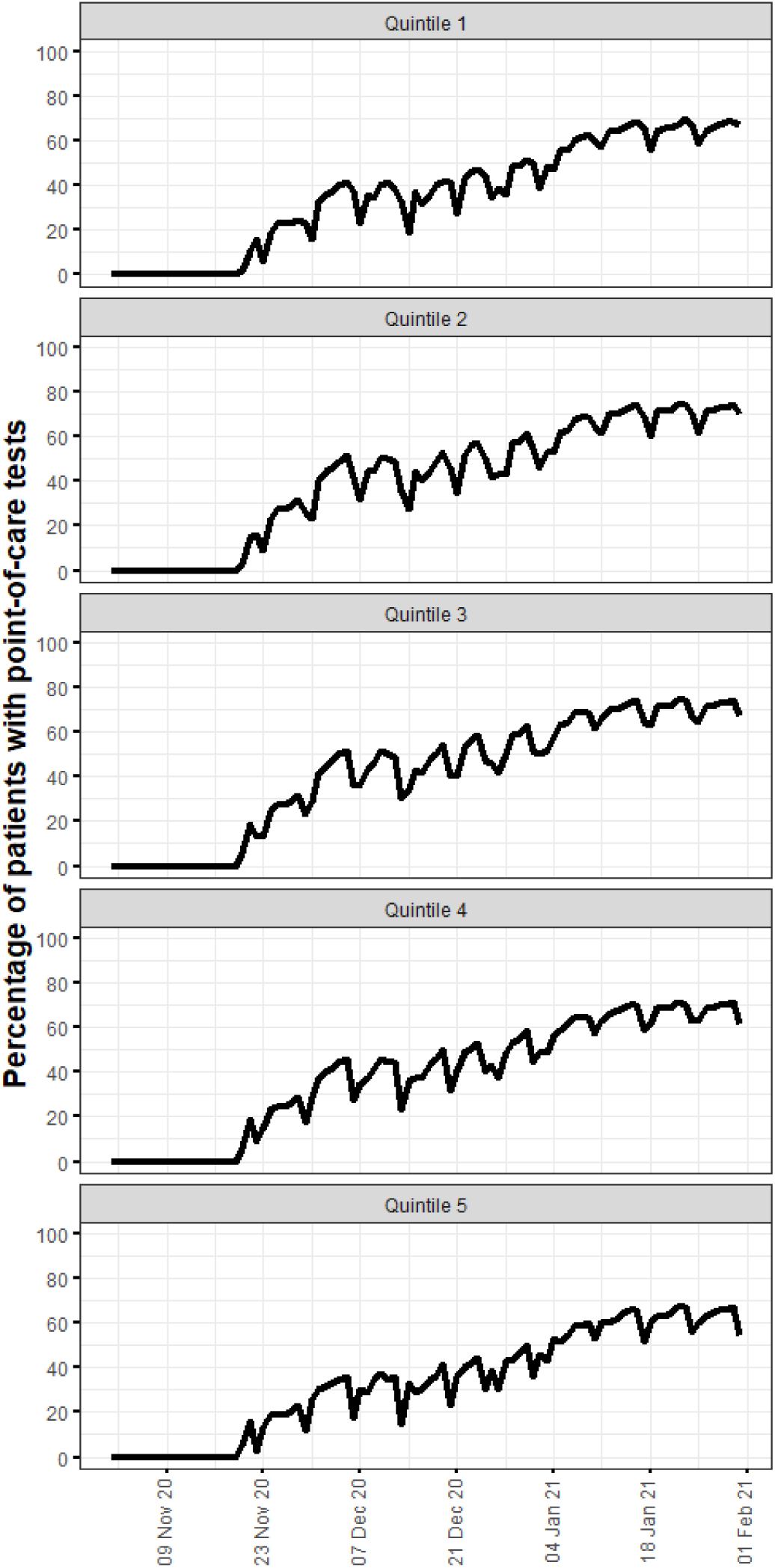
Percentage of tests on a given day that were point-of-care stratified by margination status quintile in Mexico (7-day rolling means). Quintile five represents the worst margination status.

Figures for the moving mean of latency one, latency two and total latency stratified by state, hospitalization status (only available for Mexico), and margination index are shown in Appendix Figures 2-5. Eliminating the time lag between testing and results by applying point-of-care tests would have saved a total of 2,652,774 days in Colombia (8.1 per person) and 22,172,666 in Mexico (6.9 per person) from testing to result.

## Discussion

The pandemic has caused a delay in healthcare provision in patients suffering non-covid related illnesses (*16*), but delays in patients with COVID-19 symptoms have not been thoroughly described. The time between symptoms onset and the first medical visit and sampling for tests could be determined by multiple factors such as symptoms severity, individual seeking health behavior, geographic access, and socioeconomic status, among others. After sample collection, test turnaround might depend also on geographic access but we assume that mainly on laboratory capacity and administrative procedures. Delays at each timepoint might be corrected or improved with specific strategies aiming to optimize the use of SARS-CoV-2 screening tests results for clinical case identification and isolation during the estimated period of infectiousness. We used this framework to describe the length of time between symptoms onset, testing and sample processing, and test result report for SARS-CoV-2 and its association with socioeconomic status in Colombia and Mexico. We observed that the median time between symptoms onset and tests results report exceeds that of infectiousness in both countries. There is an overall poor use of resources that depends to a great extent in a delay in tests results reporting after sample collection (*latency two* period). Most people were early testers, more so in Colombia than in Mexico, with considerable heterogeneity according to state. This supports the fact that testing strategies are not “one fit all”, and should be tailored according to each regions requirement. The presence of “local epidemics” as a way of expressing how different geographic areas are impacted differently, has been previously described (*5, 17, 18*).

Interestingly we found longer *latency two* in areas with less margination in Mexico, but longer in Colombia. This could be due to the higher use of POC tests in Mexico, and thus we are only observing RT-PCR tested patients, as well as the sole use of RT-PCR tests in Colombia during the selected study period. Since *latency one* was short and remained constant over time and across margination status, interventions to increase access to tests across the whole population, while important to test more people, are less likely to have an impact in reducing delays in the diagnostic process, as the main component of the delay is the prolonged *latency two*.

As Mexico started using antigen tests during the late second half of 2020, we explored if these were being performed preferably in vulnerable areas. Even if by the end of the study period the percentage of POC tests was similar between margination quintiles, the number of tests varied greatly, as only 18.4% of all POC tests were performed in the lower two quintiles. This is considering a similar number of people was tested among all quantiles. Thus, this reveals under testing in those most vulnerable and an unequal distribution of a valuable resource. This adds another layer to the mortality disparity seen among private and public healthcare systems in Mexico (*19*).

Pont-of-care antigen tests have been proposed as an effective strategy for epidemiologic surveillance. While their sensitivity is lower than RT-PCR tests (83-93% according to the Institute of Diagnosis and Epidemiological Reference in Mexico), their results are immediate, and their low cost easily permits repeated testing (*20*). Repeated testing of antigen tests has been shown to increase sensitivity in comparison with a one-time RT-PCR (*21*). An important argument against antigen tests is their lower sensitivity compared to RT-PCR tests. Proposed strategies based on antigen tests include repeated testing due to their low cost. For argument’s sake, even if we considered this 10% of patients as “lost” (which would imply no repeat testing) these patients would sum 1,787,732 infectious days (considering an average of 12 infectious days per person), and with the current strategy a total of 5,633,261 infectious per days are undetected solely because of latency two, three times as much.

Our study has several limitations. It is observational and used repurposed data, and as such it is difficult to ascertain the precision of initial symptom date and diagnosis date, but both countries use this data to make public health decisions and thus our results are applicable. The fact that Colombia does not share information on every tested individual is also a limitation, as positive individuals in that country could arrive at a different time to testing, even if our exploratory analysis in Mexico does not show this. Selection bias might also present, since both countries do only limited testing, with Mexico testing only one of 10 ambulatory patients and all hospitalized. We are unable to make conclusions regarding symptomatic patients who do not search for care and asymptomatic individuals, as they are not included in the open data (*22*). This could evidently influence either people’s ability to get tested and the laboratory’s speed to give results, but this is precisely our point. These variables are not accounted for in current testing strategies at both of these countries, and low testing efficiency is a side effect of this.

Our study also has several strengths. It conveys information of two countries and a large number of individuals during a significant proportion of the pandemic. Even if data is repurposed, the main variables should not have recall bias affect them in a meaningful way as it is routine information. Thus, diagnostic delays can be adequately quantified. Also, our use of DAGs makes our thought process transparent on estimating variable effect on testing delays.

The low efficiency of RT-PCR observed in our study supports points to the need of improve the efficiency of sample processing and test results reporting. Antigen tests for epidemiological COVID-19 surveillance might contribute to reduce the time between sample collection and test result delivery. Our results also indicate that efforts and resources should be more heavily invested in high margination areas and populations, which would make resource allocation more efficient.

## Data Availability

Data used in the analysis is freely available in respective government websites and code used in the analysis will be freely available with the final version of the article.

## Acknowledgments

We would like to thank the health services worldwide for their amazing effort to overcome this pandemic. Also, the data provided by Our World in Data is invaluable, and their initiative should be applauded. Finally, we would like to thank the health care systems of Mexico and Colombia, who have provided incredibly detailed open data in a true example of transparency.

