## Appendix Figure for "Delay of molecular SARS-CoV-2 testing and turnaround time in Mexico and Colombia"

**Short Title:** Delay of COVID-19 testing in Mexico and Colombia

**Authors:**

Isaac Núñez MD ^1^

Pablo F. Belaunzarán-Zamudio MD ^2, 3^

Yanink Caro-Vega PhD ^2^

**Affiliations:**

1.	Department of Medical Education, Instituto Nacional de Ciencias Médicas y Nutrición Salvador Zubirán, Mexico City, Mexico.

2.	Department of Infectious Diseases, Instituto Nacional de Ciencias Médicas y Nutrición Salvador Zubirán, Mexico City, Mexico.

3.	Department of AIDS research, National Institute of Allergy & Infectious Diseases, National Institutes of Health, Bethesda, Maryland, United States.

**Corresponding author:**

Isaac Núñez MD. Instituto Nacional de Ciencias Médicas y Nutrición Salvador Zubirán, Vasco de Quiroga #15, Tlalpan Mexico City, Mexico, postal code 14080. Telephone number: 55 5487 0900.

**Figure 1. Directed acyclic graphs for estimation of margination index/multidimensional poverty index’ effect on latency one and latency two**

**A)**

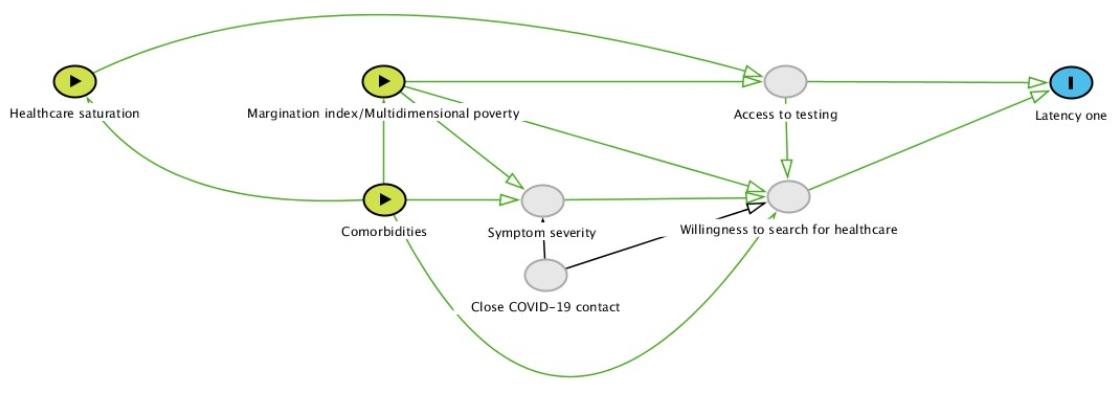

**B)**

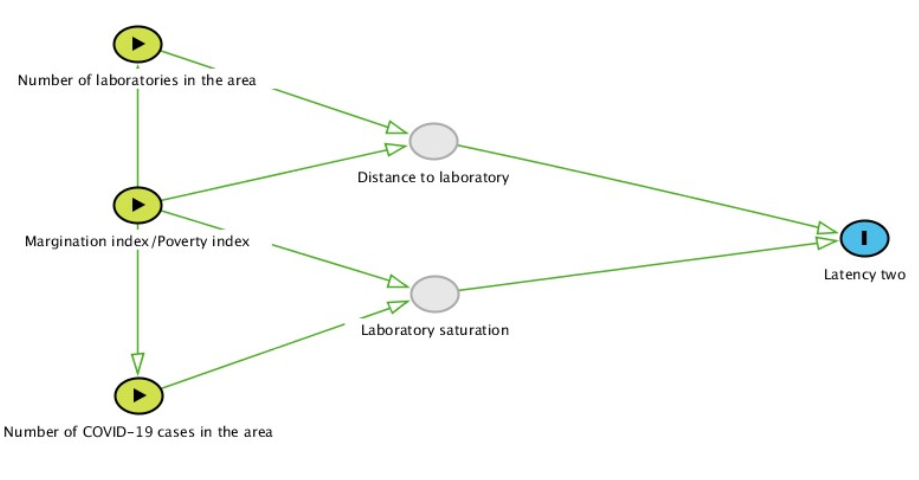

A) DAG for latency one, B) DAG for latency two. Green nodes represent observed exposures, gray nodes represent unobserved exposures, blue nodes represent outcomes.

**Figure 2. Latency periods of RT-PCR for SARS-CoV-2 in Colombia and Mexico (by multidimensional poverty index/margination index)**

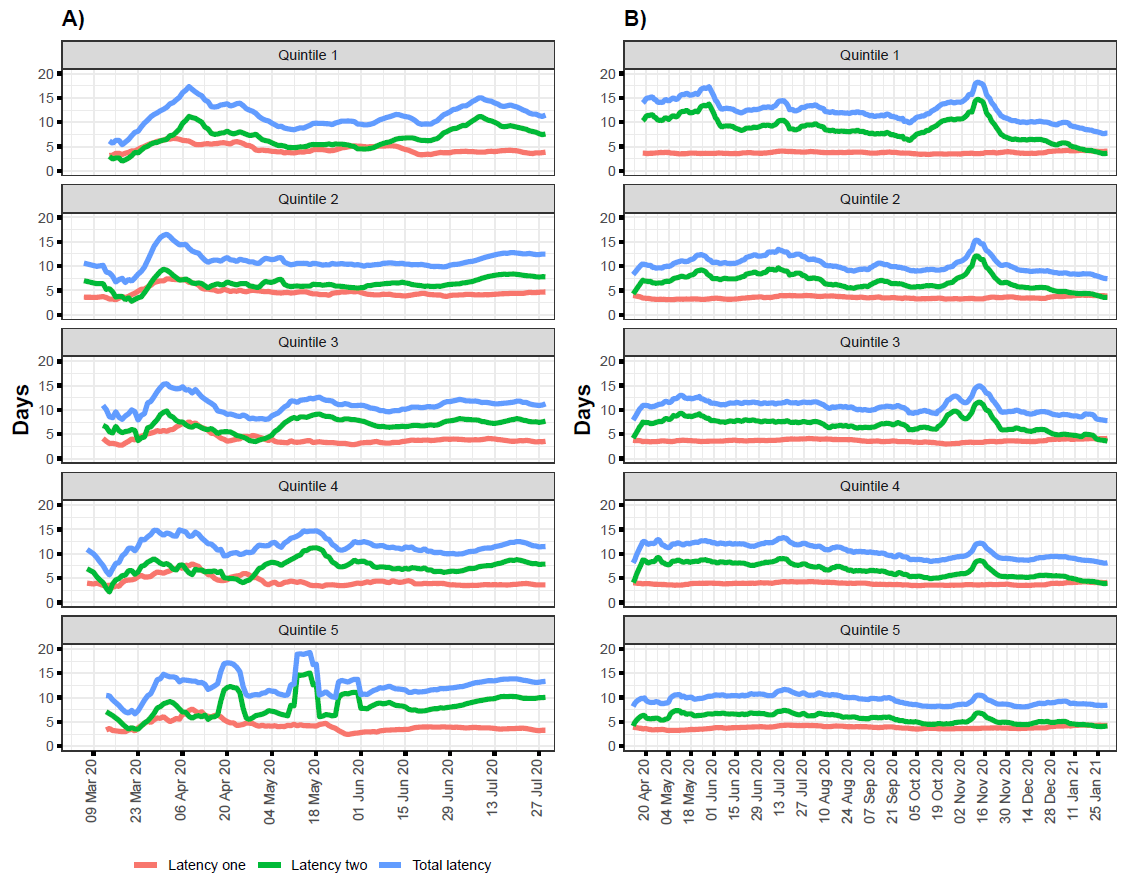

Latency one: time from first symptom until testing; Latency two: time from testing until reporting of result; Total latency: total time from first symptom until reporting of test result. Colombia uses the multidimensional poverty index, while Mexico uses the margination index. First quintile represents best status while fifth quintile represents the worst. Guainia, Vaupes, and Vichada (Colombian states) had too few observations to calculate moving means and thus were also excluded from this analysis.
